# Geographical distribution of scrub typhus and risk of *Orientia tsutsugamushi* infection in Indonesia: Evidence mapping

**DOI:** 10.1101/2023.05.26.23290567

**Authors:** K Saraswati, I Elliott, NPJ Day, JK Baird, SD Blacksell, Ristiyanto, CL Moyes

## Abstract

**Background:** Scrub typhus is a potentially fatal acute febrile illness caused by bacteria in the genus *Orientia.* Though cases have been documented, a comprehensive body of evidence has not previously been compiled to give an overview of scrub typhus in Indonesia. This study aimed to address this key knowledge gap by mapping and ranking geographic areas based on existing data on the presence or absence of the pathogen in humans, vectors, and host animals.

**Methodology/Principal Findings:** We performed searches on local and international electronic databases, websites, libraries, and collections including Embase, Medline, and Scopus to gather relevant evidence (including grey literature). After extracting data on the presence and absence of the pathogen and its vectors, we ranked the evidence based on the certainty for the presence of human infection risk. The country was divided into subnational units, and each were assigned a score based on the evidence available for that unit. We presented this in an evidence map. *Orientia tsutsugamushi* presence has been identified on all the main islands (Sumatra, Java, Borneo, Celebes, Papua). About two thirds of the data points were collected before 1946. South Sumatra and Biak had the strongest evidence for sustaining infectious vectors. There was only one laboratory confirmed case in a human identified but 2,780 probable cases were documented. The most common vector was *Leptotrombidium deliense*.

**Conclusions/Significance:** Our review highlights the concerning lack of data on scrub typhus in Indonesia, the fourth most populous country in the world. The presence of seropositive samples, infected vectors and rodents confirm *O. tsutsugamushi* is widespread in Indonesia and likely to be causing significant morbidity and mortality. There is an urgent need to increase surveillance to better understand the burden of the disease across the archipelago and to inform national empirical fever treatment guidelines.

**Author summary:** Scrub typhus is a febrile illness common in the Asia Pacific area. It is caused by bacteria in the genus *Orientia* and spread via the bite of Trombiculid mites. While we know that scrub typhus is present in Indonesia, there is very limited information on how widespread the problem is. To address this problem, this study aimed to create a map of geographic areas that are at risk of scrub typhus by analysing existing data on human, mites, and other animals. By doing so, we hope to provide a better understanding of the extent and distribution of scrub typhus in Indonesia. Evidence of *Orientia tsutsugamushi*, has been reported from all the five main islands of Indonesia. South Sumatra and Biak had the strongest evidence. However, there was no evidence of presence or absence in about two thirds of the regions and most of the data points were collected before 1946. While the available data suggest that scrub typhus is prevalent in Indonesia, up-to-date information is lacking. Therefore, it is crucial to increase scrub typhus surveillance to improve our understanding of its burden and distribution, and eventually inform treatment strategies.

## Introduction

Scrub typhus is a mite-borne acute febrile illness caused by bacteria in the genus *Orientia*. In South and Southeast Asia, scrub typhus is one of the leading causes of non-malarial fever [1]. Based on limited data, it is estimated that there are approximately one million cases annually and one billion people at risk [2].

Despite its prevalence, scrub typhus is under-recognised and understudied. The difficulty differentiating scrub typhus from other endemic febrile illnesses with similar clinical presentation, along with poor diagnostics, largely explains our limited knowledge of the burden and distribution of this disease. This uncertainty occurs even within the classical distribution of scrub typhus in Asia and the western Pacific.

There are more than 17,000 islands in Indonesia, straddled over five thousand kilometres of equator [3]. With 270 million residents, Indonesia is the fourth most populous country in the world [4]. Documentation of scrub typhus outbreaks in Indonesia exists, most notably among plantation workers in east Sumatra in the early 1900s and among Allied Forces in the western Pacific during World War II [5]. Although sparse and dated, evidence suggests that scrub typhus may be broadly prevalent in Indonesia and, as it invariably does elsewhere, causes significant morbidity and mortality. However, in the absence of a reliable overview, the burden and distribution of disease caused by scrub typhus in Indonesia remains almost wholly unknown. This paucity of information drives a vicious cycle of underrecognition, inattention, and underfunding of research that may conceal a problem of potentially great clinical and public health importance.

The objective of this work is to address these issues by categorising specific geographical regions based on evidence of their ability to harbour infectious vectors of scrub typhus or their potential to do so. This yields an evidence-based map that depicts the known geographical distribution of infection risk, which will in turn inform the development of national surveillance, diagnostic, and treatment strategies. We go on to identify specific evidence gaps for either the presence or absence of the pathogen and its vectors in Indonesia.

## Methods

### Search strategy

Thirteen international electronic databases, including Embase, Global Health, Medline, Scopus, and Index Medicus for South-East Asia Region (IMSEAR), three Indonesian electronic databases, four local university/research centre websites, five Dutch journal and museum websites, and literature held at the Eijkman Institute library were searched (Supporting Information 1).

Searches were undertaken in English and Dutch. No Indonesian language search terms were used because there is no Indonesian word for scrub typhus and pilot searches showed that Indonesian articles can be captured using Latin, English, and Dutch terms. No language or publication date restrictions were applied. The search terms were combinations of terms relating to the bacterium (*Orientia tsutsugamushi*), scrub typhus disease, e.g. mite typhus; the mite vector, e.g. *Leptotrombidium*; and alternative names for Indonesia, e.g. Dutch East Indies. The detailed search strategy is provided in (Supporting information 1). If available, we searched the reference list of relevant and review articles.

In addition, we included data from the collection of mite vector samples in the Balai Besar Litbang Vektor dan Reservoir Penyakit (B2P2VRP) – the Institute for Vector and Reservoir Control Research and Development, National Institute of Health Research Development (NIHRD), Ministry of Health (Supporting information 2). These mites were collected as ectoparasites from small mammal trapping. Finally, the results from a serosurvey we performed on human samples from Bali and Sumatra were also included [6].

### Eligibility criteria

Evidence of *O. tsutsugamushi* detection or antibodies against *O. tsutsugamushi* in humans, animals or vectors, and the presence or absence of vectors, was eligible for inclusion. Data of interest included the location of infection and details of the methods used to detect *O. tsutsugamushi* or antibodies against it. A sample size of at least 400 was required to support absence of scrub typhus. If a survey found no positive samples and its sample size was 400, using binomial distribution confidence interval calculation, the upper limit of 95% confidence interval would be 0.009, yielding 95% certainty that the true proportion of positives in the population is less than 1%.

### Data extraction

Data were extracted into a Microsoft Excel spreadsheet (Supporting Information 2). Each data point contained evidence for the presence or absence of *O. tsutsugamushi* and/or its vector at a specific location during a specified year. The location used as the data point was the presumed infection location i.e. where the patient was located for two weeks (14 days) preceding the onset of clinical signs and symptoms. When available, the number of human cases found in each data point were also collected.

In case reports or case series, the admission or first presenting date was counted as the study start date. The last day of observation or the date of discharge were entered as study end date. Where data were aggregated by location or time and could not be disaggregated, the aggregate data was entered and recorded accordingly. If the data could be disaggregated, it was separated into the smallest location or time unit possible. In the case of reinfection, each infection episode was inputted as a data point. Where the study month and year were unknown, the publication month and year were inputted instead.

### Data geolocation

The latitude and longitude of each data point were recorded in decimal format. Each data point was classed as a point or polygon. A point is defined as a site that can be geopositioned to an accuracy of ≤25km^2^ (i.e. 5km x 5km) or has ≤25km^2^ area size [7]. A polygon is a site that cannot be geopositioned to an accuracy of ≤25km^2^ or has >25km^2^ area size [7]. The coordinates of the approximate centre of the polygon were used. If the survey site coordinate information was not given or could not be located using an online gazetteer, the administrative division (e.g. subdistrict or district) was entered as the site location. The extracted data was mapped by geographic coordinates to each subnational unit.

### Data analysis

These data were synthesised using descriptive statistics, graphs, maps, and narrative synthesis. Proportions and frequencies were used to summarise and describe categorical variables. The analysis was done on Microsoft Excel 2013, StataBE 17 (StataCorp, College Station, Tx), R version 3.6.2 and R Studio Version 1.2.5033 [8, 9].

### Defining the area of study

In the mapping process, Indonesia was divided into subnational units. Indonesia is an archipelago of thousands of islands so we used a mixed approach to define subnational units where both administrative boundaries, the size of the islands, and their distance from each other were considered [7]. The largest islands in Indonesia, e.g. Java, were divided based on the level one administrative (provincial) boundary [7]. Islands larger than 200 km^2^ and further than 25 km from the largest islands were considered as separate units [7]. If the islands were within 10 km of each other, they were categorised into the same subnational unit [7]. When smaller islands (less than 100 km^2^) were ‘more than 10 km from any other islands’, they were dismissed [7]. If there were any data points located in these smaller islands, the data points were pulled to the nearest subnational unit; consequently, the data were not lost. This resulted in 77 subnational units, including the whole of Timor Island, the eastern half of which is not Indonesian territory but the nation of Timor-Leste.

### Assigning an evidence score to each subnational unit

A score was assigned to each subnational unit based on the likelihood of the region sustaining infectious vectors. The scores reflected the evidence weighted for level of certainty for the presence of human infection risk by *O. tsutsugamushi*.

The evidence was ranked in descending score order as follows: confirmed human case (score +24), probable human case (score +22), possible human case diagnosed by Weil-Felix test (score +19), possible human case diagnosed clinically without eschar (score +18), seroprevalence of >20% in community (score +16), seroprevalence of >10% and ≤20% in the community (score +15), seroprevalence of >5% and ≤10% in the community (score +14), seroprevalence of 5% or less (but >0) in the community (score +13), confirmed infection in vector (e.g. *Leptotrombidium* spp.) (score +10), confirmed infection in non-human hosts (e.g. rodents) coupled with presence of the vector (score +8), infection in non-human hosts but the presence of the vector unknown (score +6), presence of the vector but the presence of the pathogen unknown (score +3). In addition, each data point was given a score based on the year when sample collection or observation ended (Section A, Supporting Information 3). Higher scores were given to newer studies to account for possible ecological changes over time.

Confirmed cases were defined as those diagnosed with at least one laboratory method from: bacterial culture and isolation, sequencing, indirect immunofluorescence test (IFA), indirect immunoperoxidase test (IIP), polymerase chain reaction (PCR), enzyme-linked immunosorbent assay (ELISA), or immunochromatography test (ICT). Probable cases were defined as those diagnosed when eschar was found during physical examination. An eschar is a small painless necrotic skin lesion at the site of *O. tsutsugamushi* inoculation by a mite and it is a highly specific sign of scrub typhus [2, 10]. Possible cases defined as those with: first, cases with positive OX-K antigen Weil-Felix reaction; second, cases diagnosed clinically based on clinicians’ suspicion (with no eschar found). The Weil-Felix test for scrub typhus is based on the cross-reaction of the *Proteus mirabilis* Kingsbury OX-K antigen with antibodies against *O. tsutsugamushi* [11]. This test suffers from low sensitivity and specificity but is still being utilised in some resource-limited settings [11].

*Leptotrombidium* species considered confirmed vectors [12] in this study are: *Leptotrombidium akamushi*, *Leptotrombidium arenicola*, *Leptotrombidium chiangraiensis*, *Leptotrombidium deliense*, *Leptotrombidium fletcheri*, *Leptotrombidium gaohuensis*, *Leptotrombidium imphalum*, *Leptotrombidium pallidum*, *Leptotrombidium pavlovskyi*, and *Leptotrombidium scutellare* [13].

We geographically mapped the data points and assessed the evidence obtained for each subnational unit. Where there was more than one data point in one subnational unit, the evidence ranked with a higher score was used and the lower score was disregarded. We scored each subnational unit using the decision tree shown in (Fig A, Supporting Information 3). The scores for the level of evidence and sample collection year were combined to calculate the final scores. Then a map was generated with each subnational unit coloured according to its score.

### Reporting

Although this study is not a systematic review per se, there are similarities in the methodologies and reporting standards. Therefore, the reporting of this evidence mapping followed PRISMA guidelines when applicable [14].

## Results

### Search results

The initial searches were performed on 7 November 2018, with an update search on 15 June 2021. We included 51 studies for the evidence mapping (Fig 1 and Supporting Information 2) [15–65]. We also included the results from a serosurvey we performed on archived fever study samples and data from the B2P2VRP [6].

**Fig 1.**
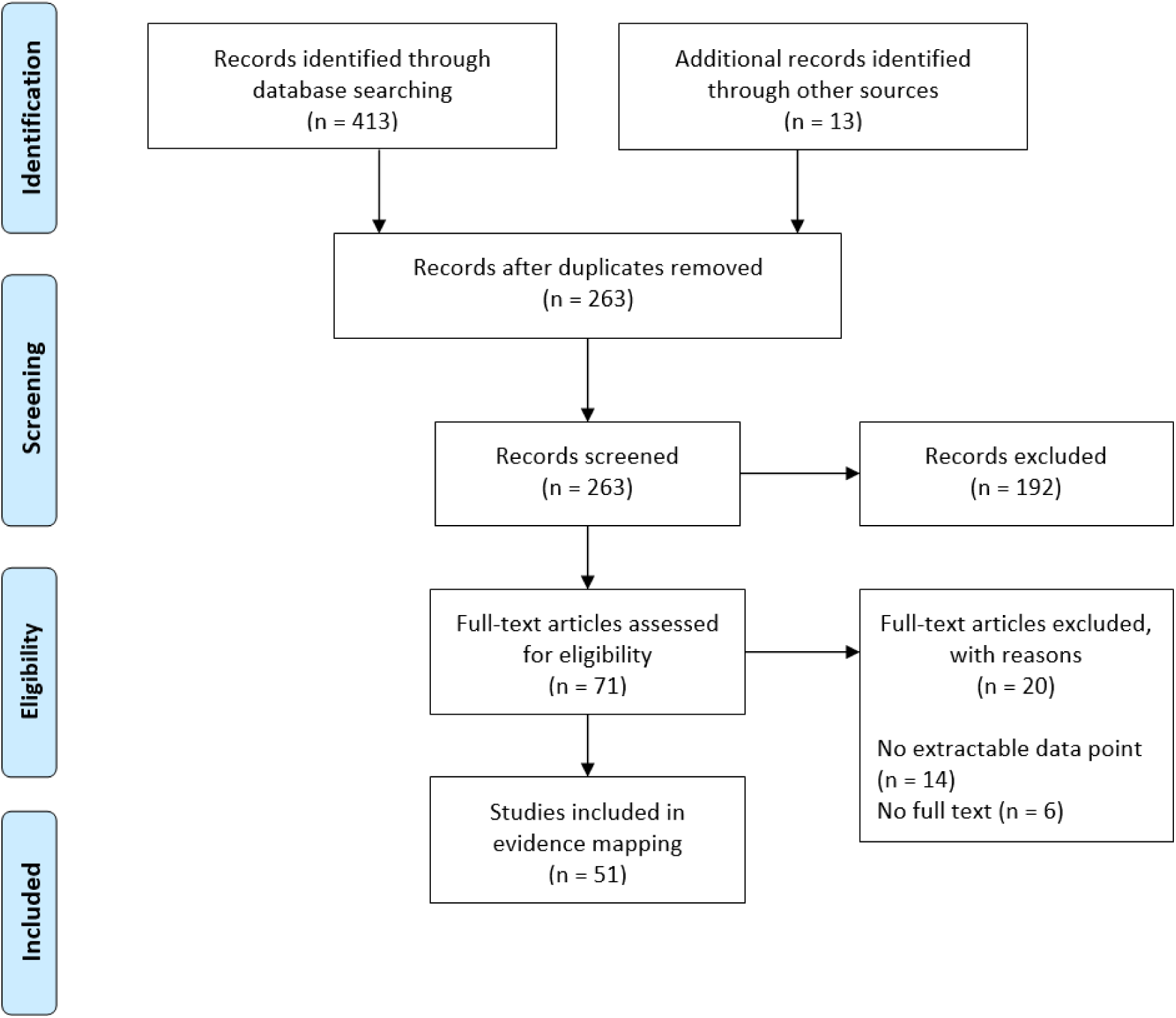
The PRISMA flowchart describing the number of articles identified in the searches, screened, assessed for eligibility, and included in the evidence mapping

### Study year

The study years range from 1908 to 2018. Most data points (n=223, 63.0%) were collected before 1946. All probable and possible human cases data points were derived from studies done prior to 1955 (Fig 2, Fig B Supporting Information 3).

**Fig 2.**
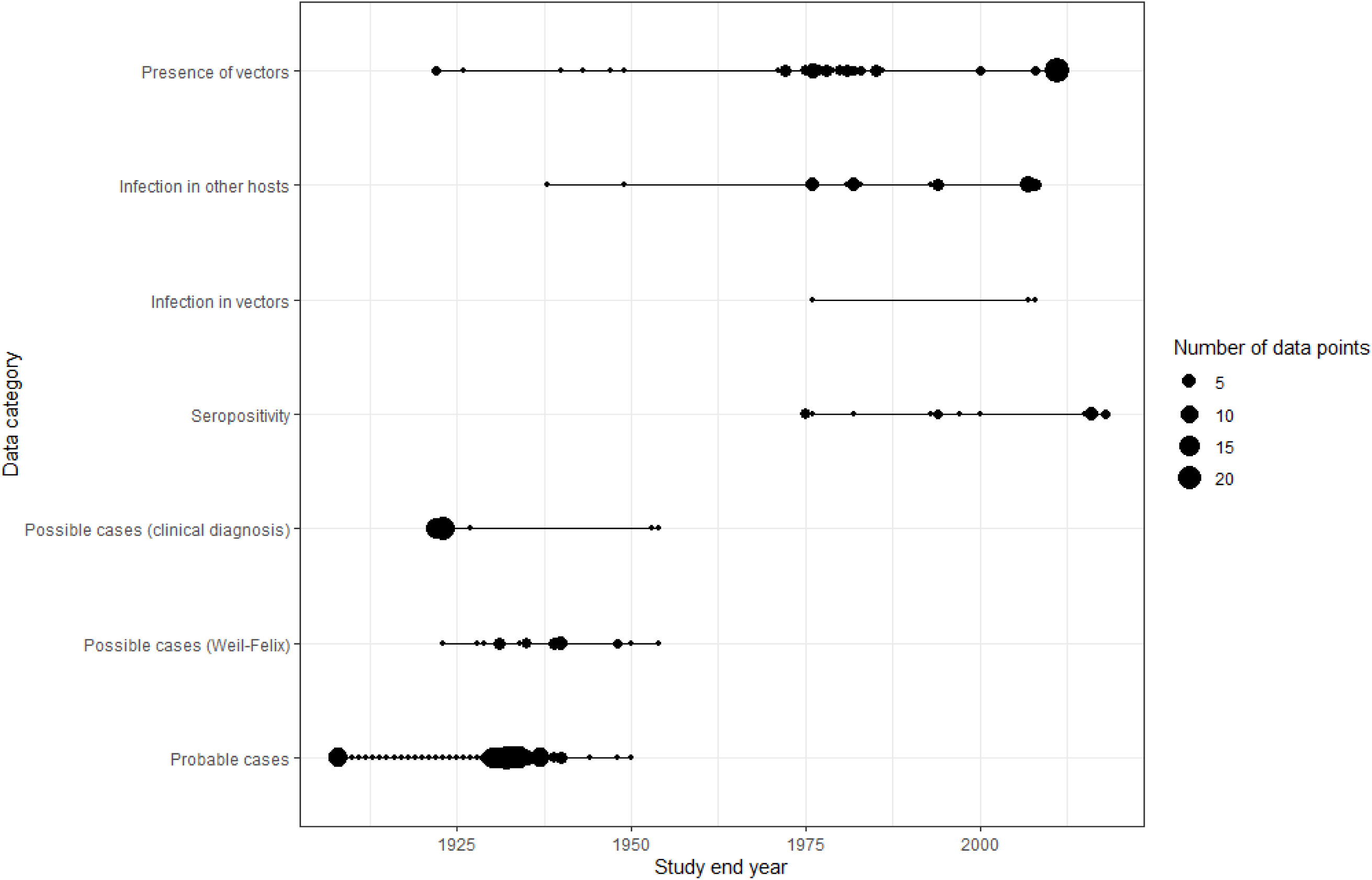
The number of data points in each category over 110 years

### Distribution of data points

There were 354 data points extracted (Supporting Information 2). These data points were distributed across 25 out of 77 (32%) subnational units, with most data points in North Sumatra (n = 149, 42.1%), followed by Aceh (n = 64, 18.1%) and Jakarta with 23 data points (6.5%) (Table 1). The median of total data points in each subnational unit was 4 (range 1–169). The three largest data point clusters were of 148 (41.8%), 50 (14.1%), 25 (7.1%) data points (Fig C, Supporting Information 3), which were in North Sumatra, Aceh, and Jakarta, respectively.

**Table 1.** The number of data points in each subnational unit

| Subnational unit | Probable case | Possible case (Weil-Felix) | Possible case (clinical diagnosis) | Sero-prevalence | Infection in vector | Infection in other hosts | Presence of vector | Number of data points |
| --- | --- | --- | --- | --- | --- | --- | --- | --- |
| North Sumatra | 103 | 3 | 39 | 0 | 0 | 0 | 4 | 149 |
| Aceh | 50 | 13 | 0 | 0 | 0 | 0 | 1 | 64 |
| Jakarta | 2 | 3 | 0 | 2 | 1 | 5 | 10 | 23 |
| East Java | 0 | 0 | 0 | 4 | 0 | 4 | 9 | 17 |
| Lampung | 0 | 0 | 0 | 4 | 0 | 6 | 6 | 16 |
| Biak | 0 | 1 | 1 | 0 | 0 | 3 | 5 | 10 |
| West Java | 0 | 0 | 0 | 1 | 0 | 0 | 9 | 10 |
| Central Java | 0 | 0 | 0 | 1 | 0 | 0 | 9 | 10 |
| North Sulawesi | 0 | 0 | 0 | 0 | 1 | 4 | 3 | 8 |
| East Borneo | 0 | 0 | 0 | 0 | 1 | 4 | 1 | 6 |
| Papua | 0 | 0 | 0 | 1 | 0 | 1 | 3 | 5 |
| West Borneo | 2 | 2 | 0 | 0 | 0 | 0 | 0 | 4 |
| Bengkulu | 0 | 1 | 0 | 0 | 0 | 0 | 3 | 4 |
| Jambi | 0 | 0 | 0 | 1 | 0 | 0 | 3 | 4 |
| Central Sulawesi | 0 | 0 | 0 | 0 | 0 | 2 | 2 | 4 |
| West Sumatra | 0 | 0 | 0 | 0 | 0 | 4 | 0 | 4 |
| West Papua | 1 | 0 | 1 | 0 | 0 | 0 | 1 | 3 |
| South Sumatra | 1 | 1 | 0 | 0 | 0 | 0 | 1 | 3 |
| Banten | 0 | 1 | 0 | 0 | 0 | 0 | 1 | 2 |
| Bali | 0 | 0 | 0 | 2 | 0 | 0 | 0 | 2 |
| South Borneo | 0 | 0 | 0 | 0 | 0 | 0 | 2 | 2 |
| Yogyakarta | 0 | 0 | 0 | 1 | 0 | 0 | 0 | 1 |
| South Sulawesi | 0 | 0 | 0 | 1 | 0 | 0 | 0 | 1 |
| Waigeo+* | 0 | 0 | 0 | 1 | 0 | 0 | 0 | 1 |
| Bintan+* | 0 | 0 | 0 | 0 | 0 | 0 | 1 | 1 |
| Total data points | 159 | 25 | 41 | 19 | 3 | 33 | 74 | 354 |

\* The notation "Waigeo+" refers to the island of Waigeo and the smaller islands surrounding it, while "Bintan+" denotes the island of Bintan and its neighbouring islets.

### Confirmed cases

A 1997 article reported an acute scrub typhus case confirmed by more than four-fold rise in ELISA IgG titre between acute and convalescent samples [66]. This patient also developed an eschar-like skin lesion. He lived in Jakarta, but travelled to the Banda Islands, Maluku within the disease incubation window [66]. Therefore, the infection site could not be pinpointed and mapped.

### Probable cases

North Sumatra had the highest number of probable case data points (n=103, 29.1%) (Table 1). There were 2,780 probable cases counted. North Sumatra had the highest number of probable case counts (n=1,651, 59.4%). Followed by West Papua with 931 probable cases (33.5%) and 34 deaths from one data point [33]. It is important to note that this number reflects the amount of testing undertaken as well as the disease incidence over a number of years and does not give an indication of infection prevalence.

### Possible cases

There were 25 data points and 158 cases diagnosed with the Weil-Felix test. Aceh had the highest number of data points (n=13/25, 52.0%) and possible cases diagnosed with the Weil-Felix test (n=77/158, 48.7%). North Sumatra had the highest number of data points (n=39/41, 95.1%) and possible cases diagnosed clinically (n = 119/147, 80.9%).

### Seropositivity

There were 11 subnational units with seroprevalence data. Lampung and East Java had the most data points (n=4/19, 21.1%). The median seroprevalence was 6% (ranging from 0.36% in Jakarta to 20% in Lampung) (Table A, Supporting Information 3). There were 3,192 people tested, with 205 found to be seropositive. The most used diagnostic methods were ELISA (n=14/19, 73.7%), followed by IFA (n=5/19, 26.3%) (Supporting Information 2).

There were three data points documenting absence, however, these data points did not fulfil our minimum sample size criteria i.e. 400. Studies in East Java and Bali both used ELISA to test 168 and 48 participants, respectively, while a study in Central Sulawesi used IFA to test 91 participants [50, 57, 65].

### Infection in vectors

There was one data point (n=1/3, 33.3%) each in Jakarta, North Sulawesi, and East Borneo. One data point recorded infection in *L. arenicola* found in Jakarta, diagnosed by animal passage and isolation [43]. Two other data points from North Sulawesi and East Borneo described infection in *Leptotrombidium* spp. confirmed with rtPCR (Supporting Information 2) [64].

### Infection in other non-human hosts

There were 33 data points for infection in other hosts. Lampung had the highest number of data points (n=6/33, 18.2%), followed by Jakarta with five data points (15.2%). All documented infected hosts were rodents (Table B, Supporting Information 3), and *Rattus tanezumi* was the most commonly found positive (n=7/33, 23.3%). Most of the data points used ELISA as the diagnostic method (n=17/33, 52%), followed by IFA (n=11, 33%), isolation (n=7, 21%), and Weil-Felix test (n=1, 3%).

### Presence of the vector

There were 74 data points reporting the presence of one or more of the confirmed vector species. Jakarta had the most data points (n=10/74, 13.5%). These data points spanned 1922 to 2011.

The most commonly found vector species was *L. deliense* (74.3%, n=55/74) followed by *L. fletcheri* (13.5%, n=10/74) (Table 2). In three data points, only the genus was identified (recorded as *Trombiculae*). These genus-only data points were not included in the main analysis since we could not be sure if they were confirmed vectors of scrub typhus.

**Table 2.** The number of data points for each vector species

| Host | Number of data point(s) | Percentage |
| --- | --- | --- |
| *Leptotrombidium deliense* | 55 | 74.3 |
| *Leptotrombidium fletcheri* | 10 | 13.5 |
| *Leptotrombidium scutellare* | 4 | 5.4 |
| *Leptotrombidium arenicola* | 2 | 2.7 |
| *Leptotrombidium akamushi* | 2 | 2.7 |
| *Leptotrombidium imphalum* | 1 | 1.4 |

### Scores

South Sumatra and Biak had the highest score for evidence for the presence of scrub typhus disease in humans with 23 points. Among subnational units with evidence, South Borneo and Bintan+ had the lowest scores with 5 points – indicating a lack of evidence (Fig 3). The median of positive scores was 18 (range 5-23). These scores were shown in the final evidence map (Fig 4). Table D, Supporting Information 3 shows the comparison between number of data points and the scores for each subnational unit.

**Fig 3.**
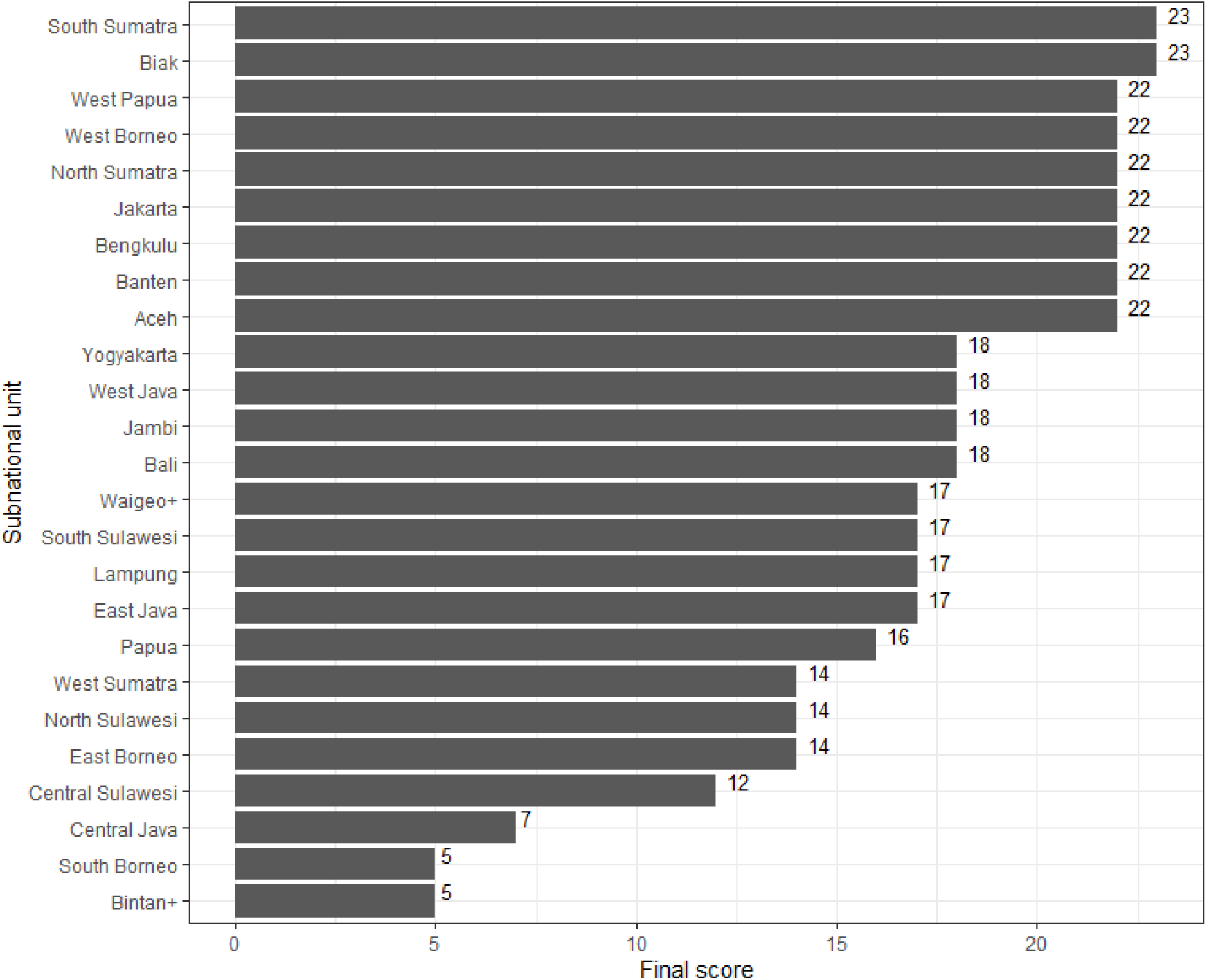
The distribution of scores across subnational units

**Fig 4.**
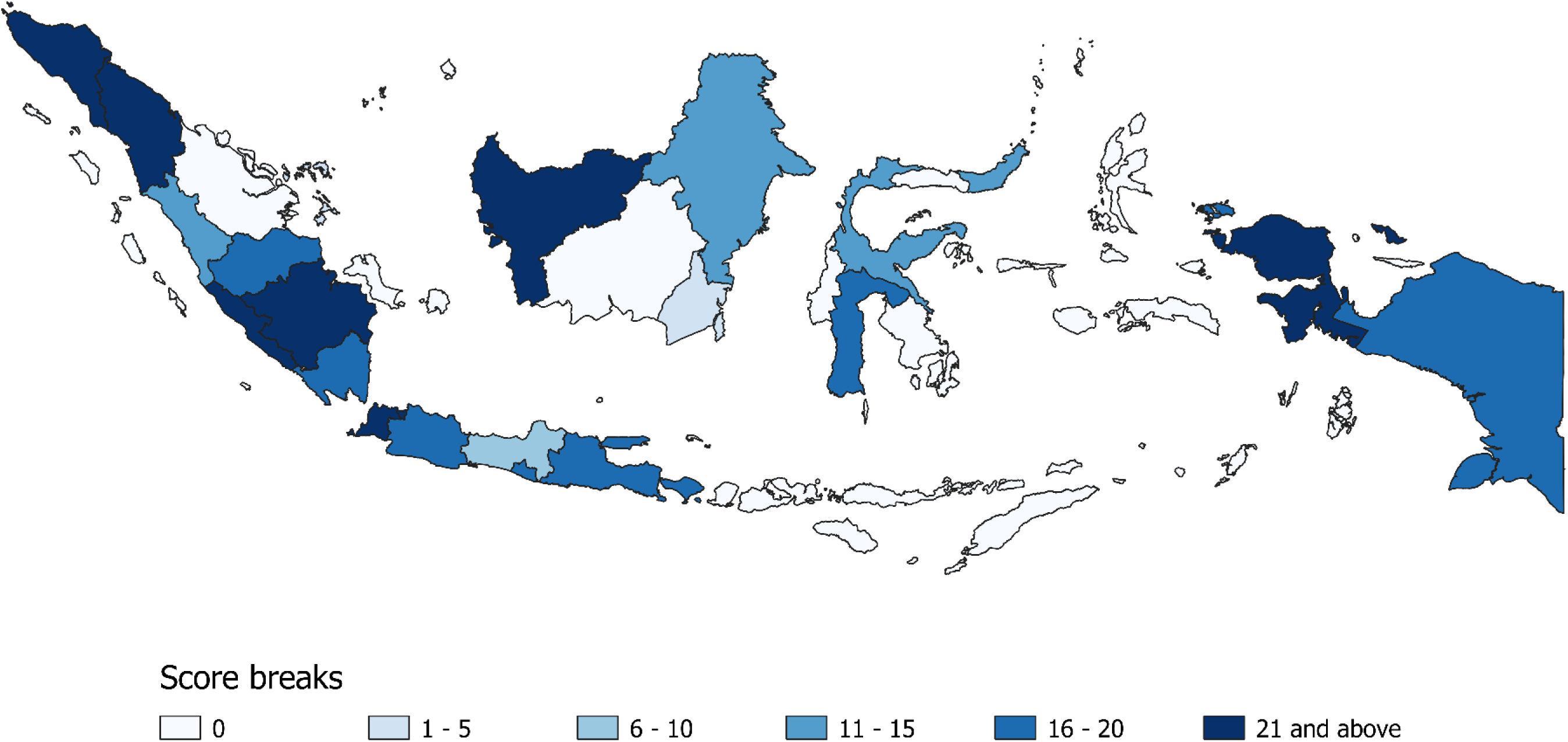
Map of evidence scores for scrub typhus / *O. tsutsugamushi* in subnational units of Indonesia

The darker the colour the stronger the evidence for presence of scrub typhus disease in humans. Low scores indicate a lack of evidence (rather than confirmed absence of this disease). Considering the range and distribution of the scores, ‘pretty breaks’ i.e. classification using round numbers as class limits, with interval of five (defined interval) were selected for visualisation [67, 68].

## Discussion

To the best of our knowledge, this is the first attempt to systematically map the evidence for the risk of scrub typhus transmission in Indonesia. Despite a lack of gold standard testing for scrub typhus in humans, we found strong evidence for the presence of *O. tsutsugamushi* infections across the Indonesian archipelago from Aceh, Sumatra in the West to Papua in the East and on all the largest islands. Within the archipelago however, we found a lack of evidence for either presence or absence of scrub typhus in two thirds of the subnational units. Further, there was a lack of large-scale epidemiological surveys that would allow areas of disease absence to be identified or infection prevalence to be estimated. South Sumatra and Biak had the best evidence available for sustaining infective vectors, but vector and pathogen surveys have not been conducted or published in many areas. This is particularly important because the widespread nature of the evidence that we did identify indicates that a risk of infection could be present in many more areas of Indonesia. Only one confirmed acute case of scrub typhus using gold standard diagnostics was documented, showing that current data on human disease is lacking [66]. However, we could not include this as a data point since the site of infection was unknown. We documented 2,780 probable human scrub typhus cases. Seropositivity data were only available in 11 out of 76 (14%) subnational units. *L. deliense* was the most common vector. We have also identified *O. tsutsugamushi* infection in *Leptotrombidium* sp. All non-human infected hosts were rodents. Almost two thirds of data points were collected before 1946.

Because the search was performed in English and Dutch only, there could be data in other languages not captured. However, this is unlikely, as publications usually mention “*Orientia tsutsugamushi*” or the term ‘tsutsugamushi’ in Latin script in the text and therefore can be captured by these search terms. To minimise the possibility of missing any relevant articles, in addition to searching in 13 international databases, we also searched the websites of three Indonesian databases, Indonesia Ministry of Health networks, three Dutch journals and three Dutch museums.

One limitation was that this study was conducted at the level of subnational regions spanning relatively large areas and a range of environments including urban and rural. The scores of each subnational unit are not necessarily reflective of the evidence available in each area within the region. This is important to keep in mind when interpreting the results, given that the distribution of scrub typhus is often described as patchy in the form of ‘mite islands’ [69].

The scores assigned to each subnational area reflect the strength of evidence for the presence of an infection risk. The scores, however, do not reflect the volume of data collected nor do they provide a value for the likelihood of infection. For areas with no evidence of presence or absence, the likelihood of sustaining infectious reservoirs is unknown. The lack of evidence for an absence of cases or infectious vectors highlights where surveys are most needed.

Almost two thirds of the data points were collected before 1946 and may no longer be representative of the current situation due to changes in land use and ecosystems [70, 71]. In addition, some of the diagnostic methods used then are not the current standard. This was mitigated in the scoring process of each subnational unit by weighting based on year of data collection, and recent evidence preferentially over older data.

This evidence mapping underlines the absence of evidence in about two thirds of subnational units in Indonesia. The lack of data points on confirmed human cases could be due to the limited availability of accurate and accessible diagnostic modalities and the low awareness of medical practitioners. Scrub typhus and other rickettsial diseases are not mentioned in the Indonesian list of medical graduates’ competencies [72]. There is an urgent need to raise awareness and include rickettsial diseases in the empiric treatment guidelines for acute fever [73]. Establishing scrub typhus and other rickettsial diseases as neglected tropical diseases recognised by the World Health Organization could increase awareness of these diseases and potentially open more research funding opportunities [74].

North Sumatra has the highest number of data points and evidence for sustaining infected vectors, however, the data points on human cases were more than 50 years old. Although the presence of infected vectors has been confirmed with rtPCR in East Borneo and North Sulawesi, we have not found any reported human scrub typhus infection or any study attempting to look for it. Regions adjoining areas with evidence of *O. tsutsugamushi* presence e.g. Riau in east Sumatra and Central Borneo, could potentially bear an unidentified scrub typhus burden. These areas should be targeted in future studies to investigate if scrub typhus is an important current public health concern.

It is possible that the incidence of scrub typhus in Indonesia is low or absent in certain areas. However, given the widespread presence of infective vectors and seropositive participants, and the presence of scrub typhus infection in nations sharing land borders with Indonesia e.g. Papua New Guinea and Malaysia [75, 76], and the current lack of survey activities and public health data, this seems unlikely. In addition to cases resembling tsutsugamushi disease reported since the 1930s, scrub typhus antibody has also been detected in Papua New Guineans [75, 77]. In Sabah (Malaysian Borneo), scrub typhus has been identified as an important but “unrecognized” cause of acute febrile illness [76].

To better understand the patchiness of infection risk within an area, species distribution models incorporating ecological parameters to predict presence in areas without data points would help fill the knowledge gaps [78]. This could also be helpful in selecting areas to do further research and direct what kind of research would be most useful. Acute febrile illness studies determining the prevalence of acute cases and serosurveys determining infection prevalence should be prioritised. Serosurveys and the inclusion of scrub typhus in the diagnostic panels for acute febrile illness studies would help lay the groundwork to better our understanding.

The available data supports a reasonable conclusion that *O. tsutsugamushi* represents a widespread infection risk in Indonesia. In some locations, the presence of vectors that can sustain infection is well documented. However, evidence mapping has also identified vast areas where data are lacking and highlights the urgent need to improve our understanding of scrub typhus across the Indonesian archipelago to inform empirical treatment guidelines for febrile patients and guide public health strategies.

## Supporting Information

S1 Appendix. Search strategy

S2 Appendix. Full dataset

S3 Appendix. Additional results

## Funding

This research was funded in whole, or in part, by the Wellcome Trust [220211]. For the purpose of Open Access, the author has applied a CC BY public copyright licence to any Author Accepted Manuscript version arising from this submission. The funders had no role in study design, data collection and analysis, decision to publish, or preparation of the manuscript.

## Data availability statement

All relevant data supporting the findings of this study are available within the article and supporting information.

## Competing interests

SDB serves as Section Editor in PLOS Neglected Tropical Diseases Editorial Board.

## Notes

### Author Declarations

Ethics approval was not required for this evidence mapping.

